# Informing an investment case for Japanese encephalitis vaccine introduction in Bangladesh

**DOI:** 10.1101/2024.03.06.24303865

**Authors:** Mariana Perez Duque, Abu Mohd Naser, Kishor K. Paul, Mahmudur Rahman, Mohammad Shafiul Alam, Hasan Mohammad Al-Amin, Mohammed Ziaur Rahman, Mohammad Enayet Hossain, Repon C Paul, Stephen P. Luby, Simon Cauchemez, Jessica Vanhomwegen, Emily S. Gurley, Henrik Salje

## Abstract

**Background:** Japanese encephalitis virus (JEV) is a major threat to human health. Bangladesh is considering introducing a JEV vaccine, however, the investment case is hampered by a poor understanding of key aspects of JEV ecology, including underlying patterns of infection, the infection fatality ratio, and which host and vectors maintain transmission.

**Methods:** We conducted a seroprevalence study in individuals of all ages in Chapai Nawabganj, Bangladesh. We tested blood samples for anti-JEV antibodies using a novel assay that limits cross-reactivity with dengue virus, trapped mosquitoes, and collected information about potential host species. We combined our results with data from a pig census, human case data and healthcare seeking patterns, all from the same region, and used mathematical models to recover risk factors for infection, and underlying probabilities of severe disease and death.

**Findings:** We found 14.3% (203/1455) of participants had antibodies against JEV. We estimated 0.5% of the susceptible population gets infected each year, however, infection risk was spatially heterogeneous, with the presence of pigs in the vicinity being the most important predictor of seropositivity. We identified 10 different known mosquito vectors for JEV. We estimated that 1 in 1,000 infections result in severe disease, 1 in 10,000 result in death, and 76% of severe cases are missed by surveillance systems.

**Interpretation:** JEV infection risk is highly spatially heterogeneous, with the underlying potential of vaccines linked to the distribution of pig-raising communities.

**Funding:** Centers for Disease Control and Prevention (CDC)

## Introduction

Japanese encephalitis (JE) is a disease caused by the Japanese encephalitis virus (JEV), a flavivirus capable of fatal encephalitis in humans. JEV was first isolated in humans in Japan in 1935, and three years later in the vector *Culex tritaeniorhynchus*^1, 2^. There are estimated to be 13,600 to 20,400 annual deaths from Japanese encephalitis globally across South and Southeast Asia ^3^. There exist effective human vaccines against JEV, with vaccine efficacies over 95% ^4^. However, not all JEV affected countries use the vaccine. The decision whether or not to use a vaccine will depend on competing health priorities, and availability of resources. Central to decision-making on whether or not to incorporate a vaccine into a national or subnational immunisation schedule is a vaccine investment case, which quantifies the potential impact of a vaccine program on human health. Bangladesh does not currently use the JEV vaccine. The first detected case of JE in Bangladesh was in 1977 ^5^, and there are cases observed each year ^6^. There is currently renewed interest in vaccinating the population.

A key complication in the development of a vaccine investment case for JEV in Bangladesh is a poor understanding of the underlying burden of infection or where risk is concentrated. In general, the disease ecology of JEV is poorly understood across the regions where it circulates, to the extent that we are still not sure of the relevance of different vectors and hosts in maintaining transmission. Humans cannot transmit JEV to mosquitoes. Instead, pigs are often cited as the main amplifying hosts, however, it has been suggested that domestic and wading birds may act as reservoirs and also play a key role in transmission^7, 8^. Similarly, a number of different vector species, including *Cx. quinquefasciatus*, *Aedes albopictus*, *Cx. vishnui, Cx. pseudovishnui*, and *Cx. tritaeniorhynchus* have been shown to be able to transmit JEV, however, their relevance to infection risk in humans remains unclear, which may also differ across transmission settings^9–11^.

There are a number of key barriers in studying JEV disease ecology. The vast majority of infections are subclinical and undetected by disease surveillance systems. Even when individuals do develop symptoms, they can be non-specific and confirmatory testing often requires cerebrospinal fluid and convalescent serum samples, which are difficult to obtain. Further, tertiary care healthcare centres that do have appropriate testing facilities are often located far from where infections occur and often not accessed, which means many severe cases remain missed^12^. Finally, serological studies that look for evidence of historic infection are complicated by the frequent cocirculation of dengue virus (DENV), another flavivirus that cross-reacts with standard commercial JEV serological assays^13^. These complications mean it has been difficult to identify risk factors for JEV infection, as well as quantify the underlying risk of severe disease and death following infection.

To fill the knowledge gap on both infection and disease burden, we focused on an area in the country where JE cases are regularly found^6^. We used complementary data sources that allowed us to explore different aspects of the disease system. Central to this effort was a seroprevalence study across individuals of all ages, where we used a novel assay that minimises cross-reactivity with DENV^14^. To capture the potential role of hosts, we used a pig census in the region that identified all pig-owning households and nomadic pig herds^8^. Further, we trapped and speciated mosquitoes and used questionnaire data to capture the presence of domestic and wading birds. Finally, we use the home location of confirmed JE cases that sought care at the local tertiary care hospital, alongside healthcare seeking studies to estimate the number of deaths and severe disease missed by surveillance systems ^12^. This series of studies from a single setting provides a unique opportunity to develop the necessary understanding on infection burden and subsequent severe disease risk to underpin an investment case when applied to the national level.

## Methods

### Data

This study brings together the results of a cross-sectional seroprevalence study, mosquito trapping study, hospitalisation data, healthcare seeking study and a pig census.

#### Cross-sectional seroprevalence study

We conducted a cross-sectional seroprevalence study in Chapai Nawabganj district between March and May in 2013. We randomly sampled 40 communities from the list of all communities in the district from the 2011 national census. In rural areas, community was defined as a village, whereas in urban areas, as a city ward. In this paper, data from 39 of these communities were used (samples from one community were not tested for JEV antibodies). Study teams visited each community, and randomly selected households for inclusion in the study. The randomisation process used the following approach: the study team first identified the house where the most recent wedding took place and then selected the closest neighbour. To select the first household of the community they counted six houses in a random direction. The last step was repeated to find the next study household. All individuals over 6 months of age were invited to participate. Following an informed consent process, individuals that agreed to participate provided a blood sample and answered a series of questions about demographic information and behaviour (e.g., travel). The household head also answered a separate questionnaire that covered characteristics of the household (e.g., household structure, animal ownership). Parents/guardians responded on behalf of individuals that were too young to answer themselves. We organised times at later dates to return to households to invite individuals from the household that were absent during the initial visit. Trained phlebotomists collected blood, which was spun down in the field. Sera was then sent to icddr,b laboratories in Dhaka in nitrogen dry shippers for serological testing. Additional households were recruited until 40 blood samples and at least 10 households had been recruited from each community. The sample size was determined based on sufficient precision to estimate the overall seroprevalence with a margin of error of under 0.05 where the true seropositivity is 0.5 (i.e. the 95% confidence interval of seropositivity would be narrower than 0.45 – 0.55). Blood group typing was provided as a benefit for all participants.

All serum samples were tested using a multiplex assay developed by the Institut Pasteur. The assay was conducted on a Luminex platform using a Bioplex 200^14^. We used a panel of beads that covered antigens from different flaviviruses (dengue serotypes 1-4, tick-borne encephalitis, yellow fever, Japanese encephalitis, and West Nile virus). Each antigen was constructed using the domain III of the E protein. This domain has been shown to have limited cross-reactivity across flaviviruses, especially as compared to commercial ELISA assays that typically use the whole E protein^14^. We considered individuals to be seropositive (i.e., had been infected with JEV) if they had a titer of 3 or greater. Luminex thresholds for JEV infection have not been defined yet, so we based this threshold on our experience with other flaviviruses (optimal cut point range of 1.5-2.1 for DENV1-4) using the same platform^15^. We conducted a sensitivity analysis with higher/lower cut points to define seropositivity (Supplementary Figure 1).

#### Mosquito trapping study

Between August and September 2015 study teams revisited the same communities as the seroprevalence study. In each community, eight randomly selected households were selected for inclusion, whether or not they had been previously included in the seroprevalence study. In households that agreed, we left a BG-Sentinel trap running for 24 hours in an indoor space. The mosquitoes were then collected and sent to icddr,b laboratories to trained entomologists for morphological identification using standard keys^16^. We recorded the number of mosquitoes by species (across males and females) trapped at each community.

#### Pig census data

We used data from a large pig census conducted between May and September 2009 through a snowball sampling technique. The results from this study have been previously published^8^. This study identified 11,364 pigs (*Sus scrofa*) in the districts of Chapai Nawabganj, alongside neighbouring Rajshahi and Naogaon districts and their coordinates were registered. We include pigs in neighbouring districts due to the potential of vector movement between districts.

#### Hospitalisation data

To help quantify the underlying level of severe disease in the region, we used data from the regional tertiary care surveillance hospital, located in Rajshahi city. This is a public hospital which receives all severe cases of referred encephalitis from the wider Rajshahi division, within which Chapai Nawabganj district is located. Confirmatory testing, consisting of both PCR and ELISA testing, was conducted at both a local laboratory and at the United States Centers for Disease Control and Prevention in Atlanta^6^. In this study, we use the year and sub-district for all confirmed JE cases that presented at Rajshahi Medical College Hospital (RMCH) between 2007 and 2016. Five out of 64 of these individuals died during the initial follow-up period (2007-2011)^17^.

#### Healthcare utilisation study

To help estimate the number of severe JE cases from Chapai Nawabganj that did not attend the hospital (RMCH), and were therefore missed by the surveillance system, we used the results of a healthcare utilisation study from the region, where study teams visited communities in the catchment area of three surveillance hospitals around the country (including RMCH) between June 2008 and March 2009. The details of this study have been published^12^. Briefly, study teams visited public meeting spaces (markets, mosques) in communities around surveillance hospitals and asked who in the community had symptoms of severe neurological disease in the past 12 months compatible with JE infection. Severe neurological disease was identified using syndromic criteria, including fever with altered mental status for more than six hours or with unconsciousness for one or more hours, or fever with altered mental status, unconsciousness, or a new onset seizure that resulted in death. They then identified the individual and interviewed them (or their family members if they had deceased) to identify the individual care-seeking pathway. From that information, we have previously quantified the probability of a severe encephalitis case being detected by the surveillance hospital as a function of the distance from the hospital^17, 18^.

### Analysis

#### Factors associated with seropositivity

We use logistic regression to identify individual-, household- and community-level factors associated with seropositivity. To account for residual spatial correlation in seroprevalence, we used a spatial random effect as implemented in INLA^19^. This was modelled as a zero-mean Gaussian process with a Matérn covariance function representing the residual spatial variation not explained by the covariates included. Due to the likely collinearity between covariates and spatial effects and to address spatial confounding in the effect estimates, we accounted for spatial dependence on covariates of interest, constraining the effect of the spatial random intercept to the outcome^20^. For that we built a design matrix with the covariates of interest and applied a QR decomposition. The resulting orthogonal matrix was then used as a constraint in the spatial random effects model. We considered models that had household and community random effects, alongside the spatial field, and decided the best model using Deviance Information Criterion (DIC). Models with DIC differences of ≤5 were considered equivalent and so we selected the more parsimonious model. We initially took all covariates in turn in univariate analysis. We then identified all covariates that were significant at the 0.05 p-value level for inclusion in a multivariable model.

#### Serocatalytic modelling

To quantify the JEV force of infection in the district, we used a serocatalytic model fit to the age-dependent variation in seropositivity^21^. The catalytic approach assumes that the risk of infection of an individual is proportional to the time of exposure (determined by their age), and therefore the probability P_p_(a) that an individual is seropositive under endemic transmission is given by the following expression ^22^:

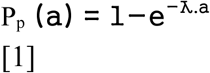

Where *a* is the age of the individual. To estimate the force of infection in each sampled community we fit the serostatus of each individual in a binomial model with a *cloglog* link function using log(age) as an offset^21^. We used INLA to approximate posterior estimates, implemented in the R package R-INLA. ^19^ To account for spatial correlation, a stochastic partial differential equation (SPDE) approach was used and the model was fitted in a Bayesian framework using INLA. We included the number of household pigs found in a 5 km radius as a covariate.

The first report of JE in Bangladesh dates from 1977^5^, however, as JEV may have been introduced earlier, we considered separate models where we assumed JEV had been first introduced at different times ranging between 1950 and 1990. To do this we adjusted the age of individuals depending on whether they were alive at the time of the first introduction. For example, when we assumed introduction was in 1977, all individuals over 38 years in age were given an age of 38. The best fitting model was determined using DIC.

From the force of infection model, we extracted the estimated force of infection in each 1km x 1km grid cell in the district. We next estimated the number of infections in each cell using the number of people living in the cell, the age distribution and the estimated level of immunity.

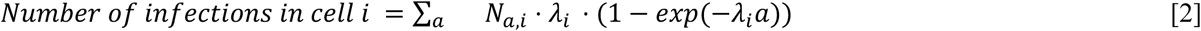

Where *N_a,i_* is the number of individuals of age *a* living in cell *i*, as estimated by WorldPop data^23^, and (1 − *exp*(−λ*_i_a*)) represents the proportion of individuals of age *a* living in cell *i* that are immune due to historic infection. This approach assumes that individuals have lifelong immunity and consequently can only get infected once.

#### Estimate disease burden using hospital-cases data

We used the results of a large healthcare-utilisation study in Bangladesh to estimate the probability that an individual with severe encephalitis would present at the Rajshahi Medical College Hospital, and therefore be detected in our human JE dataset. ^12, 17^ The methods for estimating the probability of detecting a case from the surveillance system using this data are published in detail elsewhere. ^12^ Briefly, there is a strong distance effect with individuals living farther from the surveillance hospital less likely to seek treatment there. By using the changing probability of seeking care at the surveillance hospital as a function of distance, we can estimate the number of missed cases by the surveillance system^12^. Importantly this approach is robust to underlying heterogeneity in infection risk.

#### Estimate the infection fatality ratio

The infection fatality ratio (IFR) quantifies the average number of deaths of individuals infected by a pathogen. We calculated the IFR of JEV in Chapai Nawabganj as the average number of JE deaths among all infected individuals with JEV. We estimated the IFR using the estimated average annual number of infections and the average annual number of JE deaths reported in the Chapai Nawabganj population.

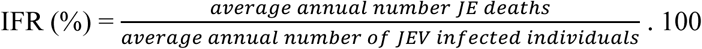

### Ethical approval

This study was approved by the ethical review boards at CDC, icddr,b, Johns Hopkins Bloomberg School of Public Health, and Institut Pasteur. Informed consent, and assent when appropriate, was obtained from all individuals.

## Results

### Human JEV infections and risk factors

A total of 1455 individuals from 393 households in 39 randomly selected communities in Chapai Nawabganj district, in Northwest Bangladesh participated in the cross-sectional seroprevalence study (Table 1). Participants had a median age of 27 years (range 2–90) and 53% were female. The age distribution of sampled individuals was largely representative of the 2011 census, except for under 5-year-olds, which were less likely to participate (Figure 1B). Overall IgG seropositivity to JEV was 19% (95% Confidence Interval (CI) 17.1-21.1), with no difference in seropositivity by sex (p-value: .6). Seropositivity increased with age, rising from 2.1% (95% CI: .7-6.2) in those <10 years to 30% in those >40 years of age (95% CI: 22.2-39.1). Mean seropositivity by community was spatially heterogeneous, ranging from 0% to 40%, with communities in the north of Chapai Nawabganj having higher seropositivity than those in the south (Figure 1A).

**Figure 1.**
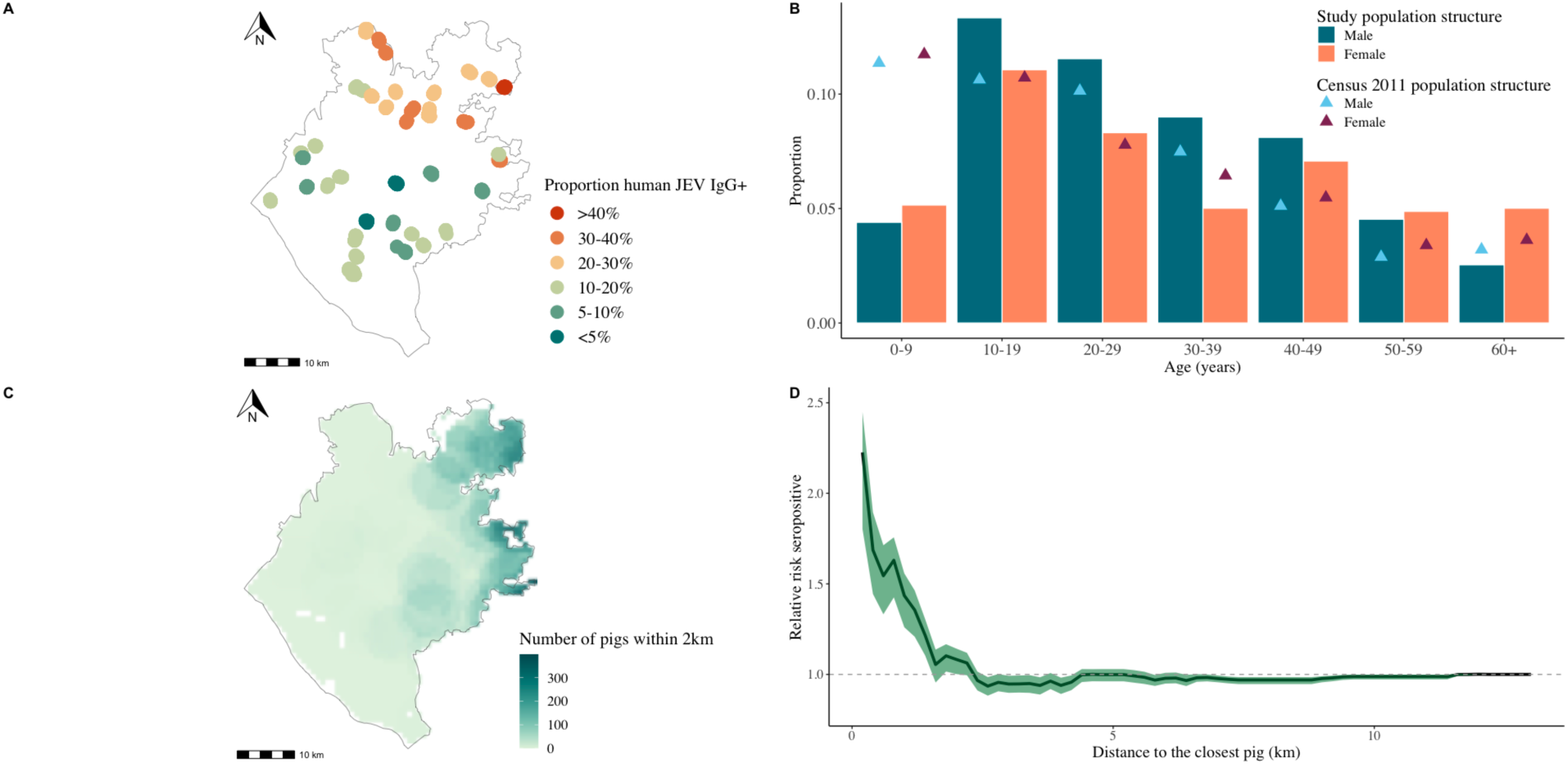
Overview of sampled communities and individuals estimates. (A) Map of sampled communities locations in Chapai Nawabganj and proportion of human individuals JEV IgG seropositive. (B) Study population structure, as proportion of total, by age group and sex (bars), and Census 2011 population structure, as proportion of total, by age groups by sex (triangles) (C) Map of sampled pigs locations (both household and nomadic). (D) Relative risk of being JEV IgG+ as a function of Euclidean distance (km) to the closest pig.

**Table 1.**
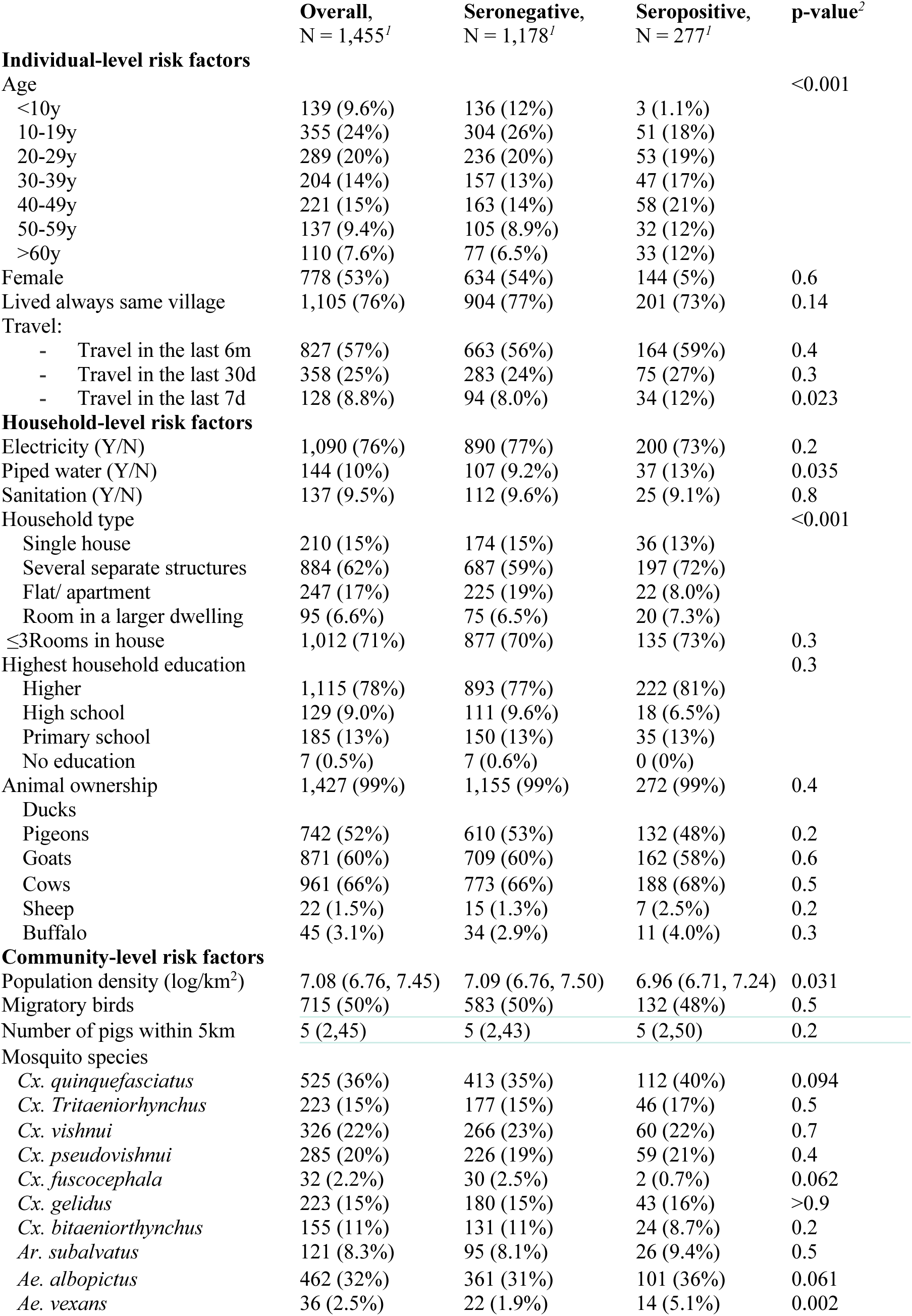
Individual-, household- and community-level characteristics of participants stratified by serostatus to JEV.

Households in four communities reported that households in their community-owned pigs. Virtually all (98%) communities had domestic duck raisers, 51% had domestic pigeon raisers, and all communities had households raising chickens. As pig ownership is relatively rare in Bangladesh, we used the results of a pig census conducted in Chapai Nawabganj to identify the distance of each individual in our study to the nearest pig owning household (Figure 1C). Individuals in our study lived on average 3km from the closest pig. Among our study participants, individuals had 1.43 (95%CI: 1.30-1.54) times the odds of being seropositive if they lived within 1km of a pig (Figure 1D). The association was attenuated when the threshold was raised to within 2km of a pig (OR 1.10, 95%CI: 1.01-1.15).

We identified ten mosquito species that have been identified as competent JEV vectors (Figure 2): *Cx. quinquefasciatus* (identified in 14/39 communities), *Ae. albopictus* (identified in 12/39 communities), *Cx. vishnui* (identified in 9/39 communities)*, Cx. pseudovishnui* (identified in 8/39 communities)*, Cx. tritaeniorhynchus* (identified in 6/39 communities), *Cx. gelidus* (identified in 6/39 communities)*, Cx. bitaeniorhynchus* (identified in 5/39 communities)*, Ar. subalvatus* (identified in 3/39 communities)*, Cx. fuscocephala* (identified in 1/39 communities), and *Ae. vexans* (identified in 1/39 communities). There was substantial spatial heterogeneity in where the different mosquito species were found (Figure 2). The vectors more commonly found in communities with seropositive individuals were *Cx. quinquefasciatus (33%), Ae. albopictus (33%), Cx. vishnui (22%)* and *Cx. pseudovishnui (19%)*.

**Figure 2.**
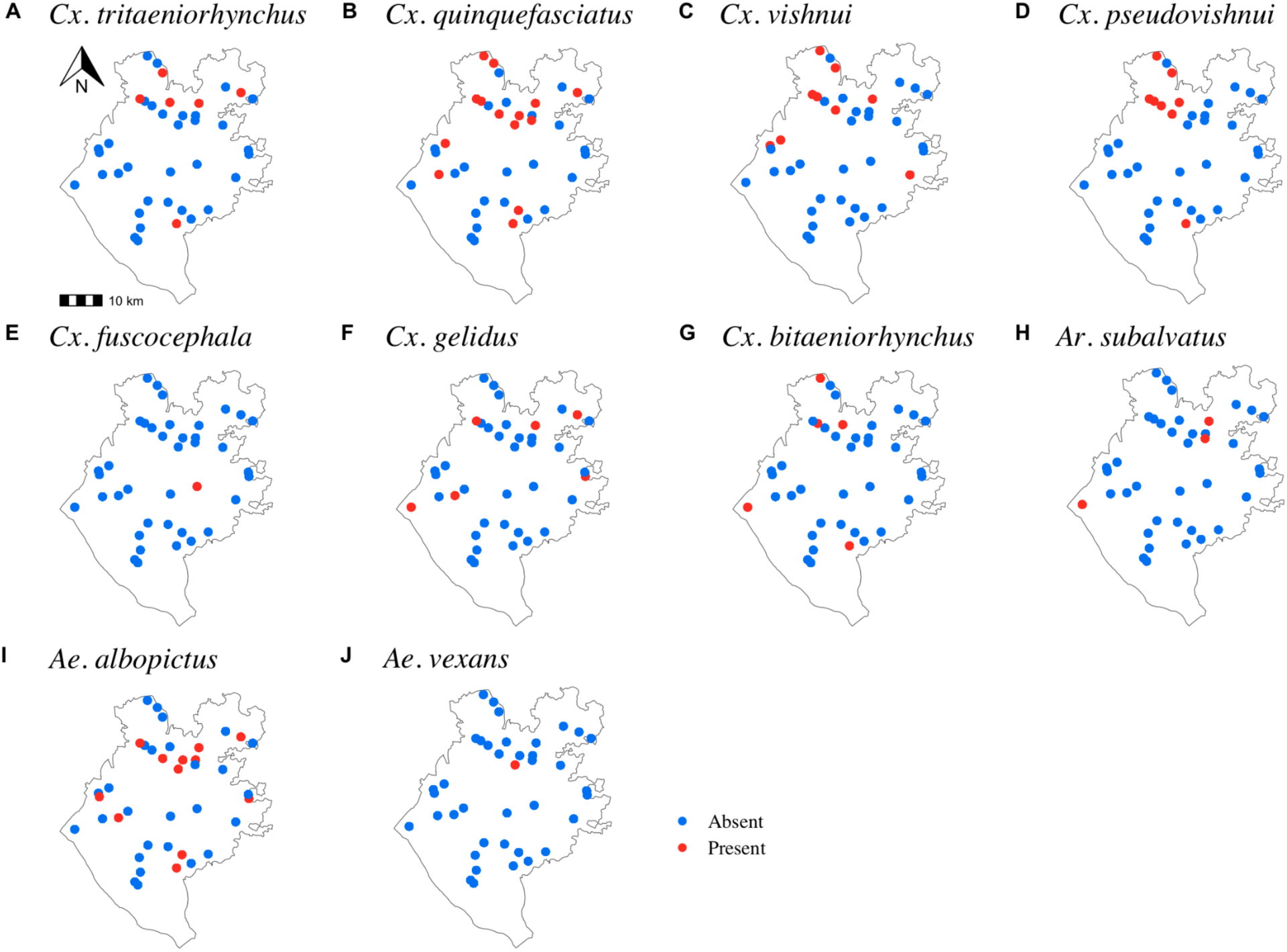
Presence of mosquito species confirmed as JEV vectors collected from mosquito traps across the district between August and September 2015.

In order to identify risk factors for being seropositive to JEV, we developed a spatially explicit logistic regression model, fit to the serostatus of each individual (Table 2). All models that included a spatial field and any combination of random intercepts had essentially the same WAIC, however, removing the spatial field resulted in a drop in model fit. We explored the relationship between different individual, household, and community risk factors in both univariate and multivariable analyses. We found the key risk factor linked to infection was the number of pigs within a 5km radius. For each 10 additional pigs within a 5km radius we found an increase of 1.02 in the odds of being seropositive (95% CrI: 1.01-1.03). Further, the presence of two vector mosquito species, *Cx*. *pseudovishnui* (aOR 1.26, 95%CrI:1.05-1.51, present in 8 communities), and *Ae. vexans* (aOR 2.03, 95%CrI: 1.41-2.92, present in a single community) were linked to seropositivity. By contrast, the location of domestic birds (ducks or pigeons) was not associated with seropositivity (Table 2).

**Table 2.**
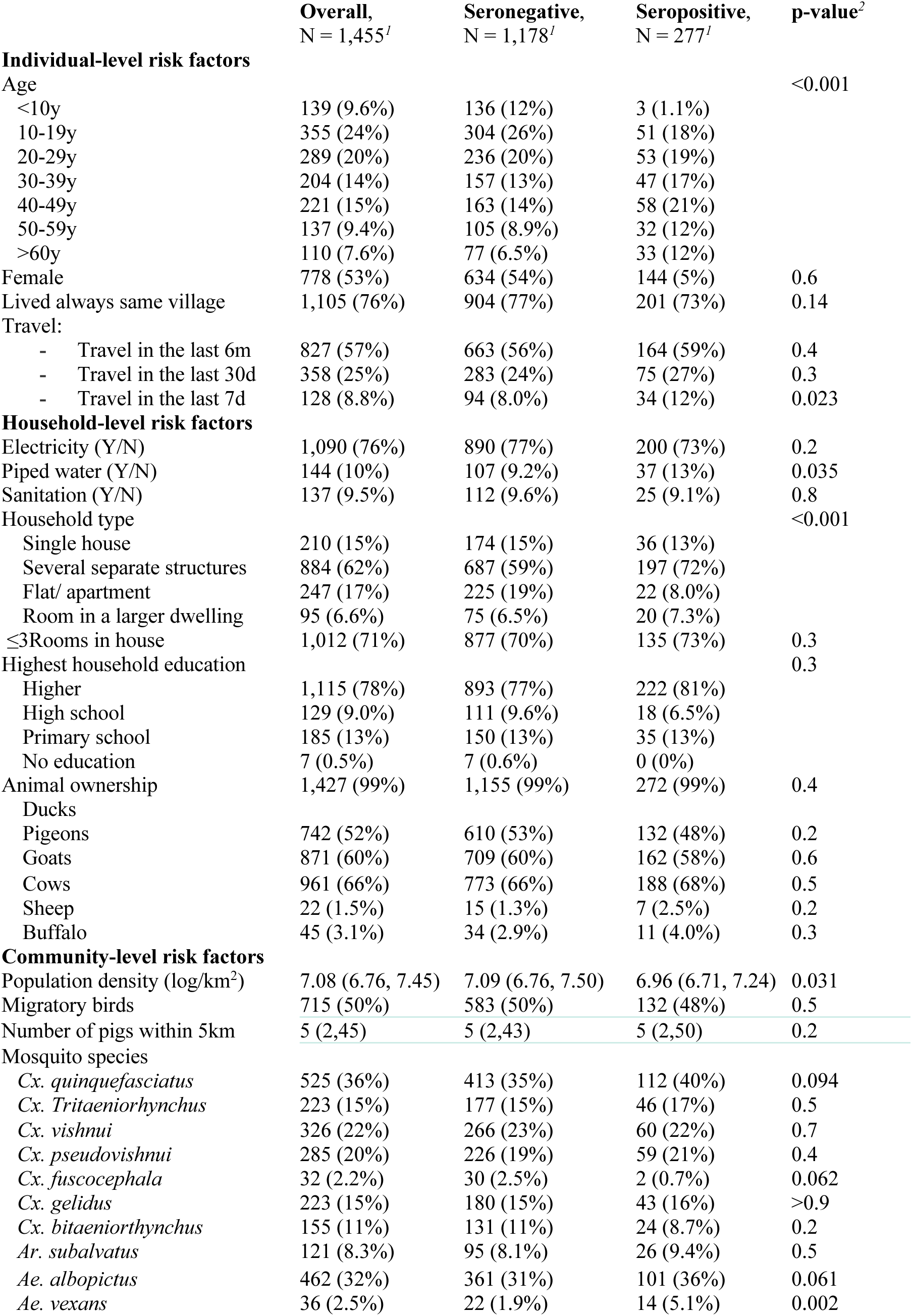
Mixed effects univariate and multivariate logistic regression model with a complementary log-log link function on the effects of individual, household and community covariates on the JEV seropositivity.

|  | Overall,<br>N = 1,455<sup>l</sup> | Seronegative,<br>N = 1,178<sup>l</sup> | Seropositive,<br>N = 277<sup>l</sup> | p-value<sup>2</sup> |
| --- | --- | --- | --- | --- |
| **Individual-level risk factors** |  |  |  |  |
| Age |  |  |  | <0.001 |
| <10y | 139 (9.6%) | 136 (12%) | 3 (1.1%) |  |
| 10-19y | 355 (24%) | 304 (26%) | 51 (18%) |  |
| 20-29y | 289 (20%) | 236 (20%) | 53 (19%) |  |
| 30-39y | 204 (14%) | 157 (13%) | 47 (17%) |  |
| 40-49y | 221 (15%) | 163 (14%) | 58 (21%) |  |
| 50-59y | 137 (9.4%) | 105 (8.9%) | 32 (12%) |  |
| >60y | 110 (7.6%) | 77 (6.5%) | 33 (12%) |  |
| Female | 778 (53%) | 634 (54%) | 144 (5%) | 0.6 |
| Lived always same village | 1,105 (76%) | 904 (77%) | 201 (73%) | 0.14 |
| Travel: |  |  |  |  |
| - Travel in the last 6m | 827 (57%) | 663 (56%) | 164 (59%) | 0.4 |
| - Travel in the last 30d | 358 (25%) | 283 (24%) | 75 (27%) | 0.3 |
| - Travel in the last 7d | 128 (8.8%) | 94 (8.0%) | 34 (12%) | 0.023 |
| **Household-level risk factors** |  |  |  |  |
| Electricity (Y/N) | 1,090 (76%) | 890 (77%) | 200 (73%) | 0.2 |
| Piped water (Y/N) | 144 (10%) | 107 (9.2%) | 37 (13%) | 0.035 |
| Sanitation (Y/N) | 137 (9.5%) | 112 (9.6%) | 25 (9.1%) | 0.8 |
| Household type |  |  |  | <0.001 |
| Single house | 210 (15%) | 174 (15%) | 36 (13%) |  |
| Several separate structures | 884 (62%) | 687 (59%) | 197 (72%) |  |
| Flat/ apartment | 247 (17%) | 225 (19%) | 22 (8.0%) |  |
| Room in a larger dwelling | 95 (6.6%) | 75 (6.5%) | 20 (7.3%) |  |
| ≤3Rooms in house | 1,012 (71%) | 877 (70%) | 135 (73%) | 0.3 |
| Highest household education |  |  |  | 0.3 |
| Higher | 1,115 (78%) | 893 (77%) | 222 (81%) |  |
| High school | 129 (9.0%) | 111 (9.6%) | 18 (6.5%) |  |
| Primary school | 185 (13%) | 150 (13%) | 35 (13%) |  |
| No education | 7 (0.5%) | 7 (0.6%) | 0 (0%) |  |
| Animal ownership | 1,427 (99%) | 1,155 (99%) | 272 (99%) | 0.4 |
| Ducks |  |  |  |  |
| Pigeons | 742 (52%) | 610 (53%) | 132 (48%) | 0.2 |
| Goats | 871 (60%) | 709 (60%) | 162 (58%) | 0.6 |
| Cows | 961 (66%) | 773 (66%) | 188 (68%) | 0.5 |
| Sheep | 22 (1.5%) | 15 (1.3%) | 7 (2.5%) | 0.2 |
| Buffalo | 45 (3.1%) | 34 (2.9%) | 11 (4.0%) | 0.3 |
| **Community-level risk factors** |  |  |  |  |
| Population density (log/km<sup>2</sup>) | 7.08 (6.76, 7.45) | 7.09 (6.76, 7.50) | 6.96 (6.71, 7.24) | 0.031 |
| Migratory birds | 715 (50%) | 583 (50%) | 132 (48%) | 0.5 |
| Number of pigs within 5km | 5 (2,45) | 5 (2,43) | 5 (2,50) | 0.2 |
| Mosquito species |  |  |  |  |
| *Cx. quinquefasciatus* | 525 (36%) | 413 (35%) | 112 (40%) | 0.094 |
| *Cx. tritaeniorhynchus* | 223 (15%) | 177 (15%) | 46 (17%) | 0.5 |
| *Cx. vishnui* | 326 (22%) | 266 (23%) | 60 (22%) | 0.7 |
| *Cx. pseudovishnui* | 285 (20%) | 226 (19%) | 59 (21%) | 0.4 |
| *Cx. fuscocephala* | 32 (2.2%) | 30 (2.5%) | 2 (0.7%) | 0.062 |
| *Cx. gelidus* | 223 (15%) | 180 (15%) | 43 (16%) | >0.9 |
| *Cx. bitaeniorhynchus* | 155 (11%) | 131 (11%) | 24 (8.7%) | 0.2 |
| *Ar. subalvatus* | 121 (8.3%) | 95 (8.1%) | 26 (9.4%) | 0.5 |
| *Ae. albopictus* | 462 (32%) | 361 (31%) | 101 (36%) | 0.061 |
| *Ae. vexans* | 36 (2.5%) | 22 (1.9%) | 14 (5.1%) | 0.002 |

### Underlying JEV infection burden

We next used our spatially explicit regression framework to estimate the infection burden from JEV throughout the district of Chapai Nawabganj. The seroprevalence model with a constant FOI over time aligns with the time of the first introduction of JEV into the region being in the late 1970s (Figure 3A) and outperforms models where we either assume JEV has been endemic since the 1950s, or with a more recent introduction (Supplementary Figure 2). We therefore used a model with a date of introduction of 1977 for subsequent analyses, consistent with the observed first introduction in the country. We used a model with distance to nearest pig (as estimated by the pig census) ^8^ and the spatial distribution of the population in the district as estimated by WorldPop data ^24^ and the yearly age distribution from the 2015 population estimates ^25^. We estimated a mean annual force of infection in the district of 0.005 (95%CI: 0.003–0.007), leading to an average seropositivity of 12.7% (95%CI: 7.0–23.3). The force of infection varied significantly over the region, with infection risk concentrated to the northwest, in the upazilas of Gomastapur and Nachole (Figure 3B). These estimates suggest that 13,178 (95%CI: 11,528 –15,062) individuals are infected each year in the district (Figure 3D). To validate the model, we used leave-one-out cross validation, where we systematically removed all data from each community from the model fitting process and compared the community-level seropositivity predicted for the held-out community with that actually observed. We found a good correlation (Pearson coefficient r= 0.63) between the predicted and observed values (Figure 3C).

**Figure 3.**
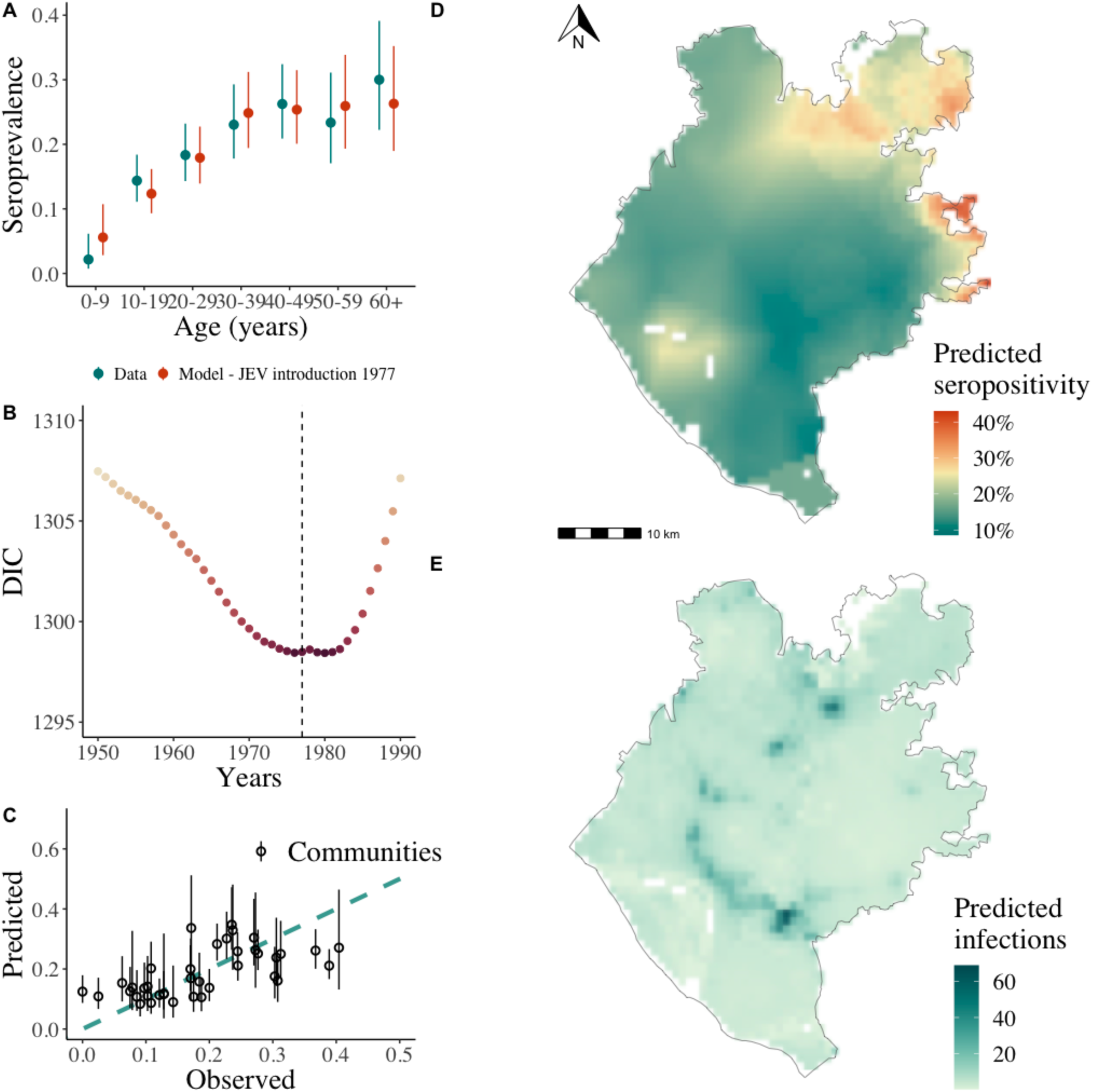
Modelled seroprevalence (A) Observed (blue points and range) and fitted seroprevalence (orange points and range - spatial model with JEV introduction in 1977) by age group for sampled individuals. (B) Deviance Information Criterion (DIC) values of each spatial hierarchical model with JEV introduction year from 1950 to 1990. Dashed line represents 1977, the year of the first case diagnosed. (C) Cross-validation using leave-one-out predictive performance of the modelled seroprevalence by community. (D) Spatial JEV seroprevalence prediction map using a Matérn covariance structure, Chapai Nawabganj. (E) JEV predicted annual infections map, 2015, Chapai Nawabganj.

### Severe case burden in humans

The Government of Bangladesh maintains a hospital-based acute meningitis-encephalitis syndrome surveillance for JE, which means that it depends on hospital attendance to detect JE severe cases. From 2007 to 2016, there were 29 confirmed Japanese encephalitis cases from Chapai Nawabganj attending the closest tertiary care surveillance hospital, located in neighbouring Rajshahi^6^. The mean age of cases was 35 years (range 0-70 years). Patients were followed up between 2007-2011 and five deaths were reported among 64 severe JE cases that presented at the surveillance hospital. There were differences in the number of JE severe cases by distance from hospital, with most cases (66%) located within 60 km of the Rajshahi surveillance hospital (Figure 4A-B).

**Figure 4.**
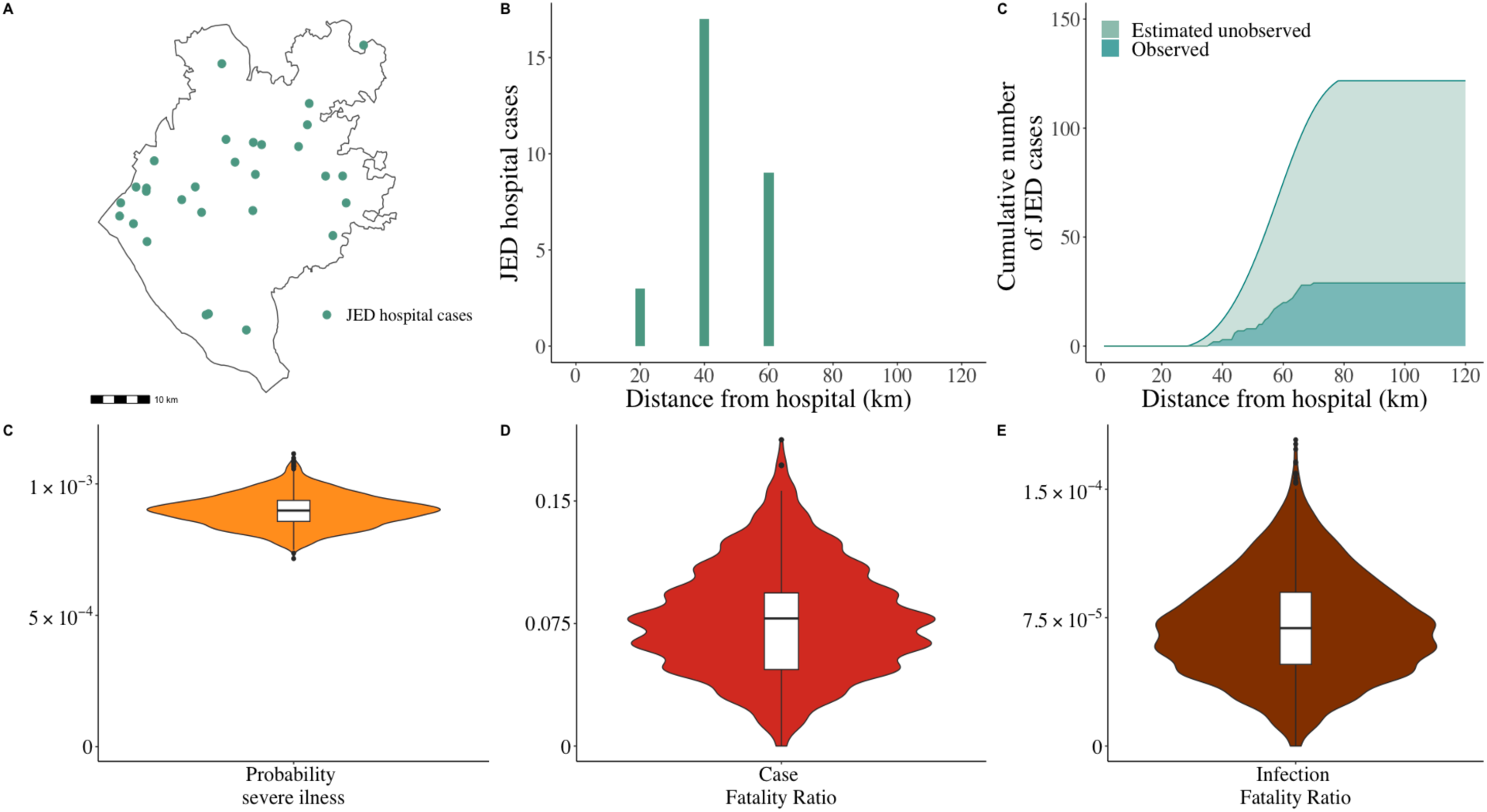
Hospital JE severe cases (A) Distance from the hospital (km) of each observed JE severe case (n=29), Chapai Nawabganj. (B) Observed JE severe cases as a function of distance (km) from the surveillance hospital. (C) Cumulative numbers of observed and estimates of unobserved JE severe cases using a healthcare seeking behaviour regression model. (D) Case Fatality Ratio of JE in Chapai Nawabganj using deaths observed in hospital and observed JE severe cases. (E) Infection Fatality Ratio of JE in Chapai Nawabganj using the probability of JE severe symptoms, obtained from total estimated JE severe cases over estimated number of infections in the district, and previously estimated cases fatality ratio.

To estimate the number of Japanese encephalitis cases that did not attend this surveillance hospital, we made use of a healthcare seeking study conducted in the region ^17^. We estimated that between 2007 and 2016 there were an additional 92 severe Japanese encephalitis cases (95% CI: 87-95) from Chapai Nawabganj that did not seek care at the surveillance hospital, resulting in 121 (95% CI: 117-125) total severe cases over this period. We estimated that there were overall nine deaths from JEV over this period (95%CI: 2-19), five of which were detected. This results in an average of 12 severe cases and one death (95%CI: 0-2) in the district per year. The resulting estimated JE incidence of severe cases in Chapai is 0.7 cases per 100,000 inhabitants. It also suggests that 24% (95%CI: 23-25) of severe JE cases are detected by the surveillance system. Comparing the number of severe cases with the estimated number of annual infections suggested that 0.09% (95%CI: 0.08-0.11) of infections result in severe disease (Figure 4C). Further, we estimated the case fatality ratio to be 7.8% (95%CI 1.6-14.1) and the infection fatality ratio (IFR) to be 0.01% (95%CI: 0.002-0.02) (Figure 4D/4E).

### Comparison of assay response to DENV

Our seropositivity results are reliant on a novel assay that uses just the EIII domains of the flaviviruses, which are more specific than the whole E protein ^14^. To assess whether the assay can specifically identify JEV infections and is not affected by DENV infections, we compared the relative fluorescence intensity to the JEV antigen with the mean relative fluorescence intensity to the four DENV1-4 antigens (Supplementary Figure 3). We find a mean correlation of 0.17 across the four serotypes (DENV1 0.07, DENV2 0.25, DENV3 0.07, DENV4 0.28). Overall seropositivity to any DENV as measured by the Luminex was 19.5 (95%CI: 17.6-21.6), and higher in the more urban south of the district, as compared to the more rural north, consistent with JEV and DENV occupying different ecological niches in this area (Supplementary Figure 4).

## Discussion

In this study we have combined a population-based serology study, a novel assay, and detailed descriptions of candidate host species as well as human case data and healthcare seeking data to obtain a detailed characterisation of the drivers of infection risk of a major but poorly understood burden on public health. We have also generated estimates of the underlying level of infection and the probability of severe disease and death following infection. Our findings confirm the endemic nature of JEV in Bangladesh and suggest JEV emergence in a previously naive population around 1977, consistent with the first detected case in the country^5^.

The estimated incidence of severe JE disease in the district (approximately 1 case per 100,000) is similar to other low endemic countries before introduction of the vaccine and lower than many highly endemic regions ^26,27^. On average under 1% of the population gets infected annually, and around 1:1,000 of infections result in severe disease and 1:10.000 result in death. Our estimates of the asymptomatic/symptomatic ratio are consistent with existing estimates^28^. Importantly, however, previously estimates have relied on ELISA assays conducted between 1950-1990, during which cross-reactivity issues with other flaviviruses were potentially encountered. ^28–30^ While low on average, infection risk was spatially heterogeneous, and was higher in the vicinity of pig-raising communities. Bangladesh is predominantly Muslim, with pig-raising households usually in small, ethnic minority rural communities.^8^ Optimal deployment of a vaccine and impact on health per dose may come from focusing on districts where risk is greatest, likely in regions with the greatest number of pig-raising households. However, subnational vaccination efforts may be difficult for national vaccine programs to implement. There also seems to be a risk of further marginalising the ethnic minorities engaged in pig raising^31^. A vaccine program focused on the pigs themselves may also be considered although the rapid turnover in pigs due to short life span and presence of maternal antibodies, which last 2-3 months, may make this unfeasible^8, 32^. While infection risk appears to have been relatively stable since JEV’s introduction in the region, the large number of competent vectors coupled with a warming climate may lead to increases in future transmission risk^33, 34^.

We identified multiple mosquito species that are known competent vectors for JEV. Some vector species, such as *Cx. pseudovishnui* and *Ar. subalvatus* were previously collected in Bangladesh mosquito surveys with high frequency, and specifically in Chapai Nawabganj ^35^. Additionally, both *Aedes* species highlighted in our analysis have been profiled in Chapai Nawabganj, nevertheless in low frequency (<5% of the collection efforts), using light traps near hosts (pigs). Although different species were associated with JEV seropositivity, *Cx. quinquefasciatus* was the most common species in communities where JEV seropositivity was observed. Previous field studies around cattle in the same region identified *Cx. tritaeniorhynchus* as the most frequently collected known JEV vector ^35^. We did not find an important role of these two vectors in our analysis, highlighting that sampling around non-human dead-end hosts, such as cattle, might not identify the vector’s role in human JEV risk. The presence of cattle in a community did not appear to be protective, as has previously been suggested, although given the overall low level of JEV transmission, we may not have had the power to detect such a protective effect ^36^. Our findings suggest that multiple mosquito species may contribute to driving infection risk in this region. We note that our traps were left in indoor spaces only, and therefore we did not capture the potential other vectors that only are found outside the household.

We identify relative proximity to pigs as the most important risk factor for human infection. While domestic and peri-domestic birds can become infected by JEV ^37–39^, they do not appear to be important in maintaining transmission in this region. Essentially all the sampled communities had a large number of domestic birds. If they were driving JEV transmission, we would observe a much more even pattern of seropositivity across the region. There were nevertheless human cases and seropositive individuals far from pig-raising households. It remains unclear whether travel to endemic regions, the presence of nomadic pig herds that travel across the wider region, the presence of other host species (e.g., wading birds) or long travel of infected mosquitoes from elsewhere could explain these infections.

Our findings confirm that the observed number of confirmed Japanese encephalitis cases by the surveillance system only represents a minority of the true number of severe cases, with around three-quarters of severe cases missed. In particular, we note that the tertiary care surveillance hospital in the region, which is relied on for confirmatory JEV testing (near the south), is farthest from the part of the region with greatest infection risk (the north). Healthcare seeking in Bangladesh is dominated by the informal sector, especially at early stages of disease^12^. Limited availability of public transport severely hampers access to tertiary care governmental healthcare facilities. Our findings support the targeted expansion of JEV surveillance systems to the north of the district. They also demonstrate the utility in combining community-based healthcare seeking surveys with surveillance data using existing platforms to obtain estimates of underlying burden ^40^. Similar approaches have been conducted to estimate the number of missed Nipah virus cases, which also circulate in the same region^18^.

Our study highlights the strength of spatial community-based seroprevalence studies to quantify the burden of infection and identify at risk populations, especially for pathogens where asymptomatic infection is common. Similar studies from the country have explored the spatial distribution of risk for multiple pathogens using the same blood samples ^41–43^. The recent growth in the use of multiplex technology, especially via the Luminex platform, alongside advances in the development of antigens that limit cross-reactivity are central to these efforts^44^. Our samples were tested against the four DENV at the same time, and the resultant titers had a correlation of around 0.2. We also found low overall seropositivity to DENV as compared to other parts of Bangladesh (e.g. Dhaka, Chittagong) and other countries in the region where DENV circulates, and importantly DENV seropositivity appeared concentrated to different locations than JEV seropositivity, consistent with the two pathogens occupying different ecological niches in the district. Further, by triangulating with disease data from surveillance, we can obtain a better understanding of the probability of severe outcomes following infection.

Our study results should be interpreted considering some limitations. Our datasets do not overlap in time. The pig census was conducted in 2009, the healthcare seeking study in 2011 and the seroprevalence study in 2015, and therefore do not capture the most recent epidemiological situation in the district. They also only represent single snapshots of the situation at the time of data collection. However, we note that the location of pig raising communities is unlikely to have changed over these timeframes, and our findings are consistent with a very stable level of annual JEV transmission since the late 1970s, which would mean any changes in inference are likely to be minor. We also predicted the burden in unsampled communities using covariates of interest. While our sampling strategy to select communities for inclusion was random, we may have missed pockets of transmission. Finally, there may be residual cross-reactivity with flavivirus circulating in the region. In particular, we estimated a substantial number of infections in the more populated southern part of the district. Even a lower level of cross reactivity with DENV could explain these infections, which would mean true JEV burden may be slightly lower than our estimates. Improved assays or analytical approaches may improve the resolution of the test results.

Despite these limitations this study presents a comprehensive assessment of an important and understudied burden on health, identifies risk factors for infection and quantifies the probability of severe disease and death. Our methodology framework applied to national level data on JE severe disease cases, national representative seroprevalence studies and risk factors exposure could have direct implications for the development of investment cases for a vaccine introduction at a country level by identifying core areas for vaccine introduction.

## Data Availability

Data and source code to reproduce analyses will be available at https://github.com/marianaperezduque.

## Contributors

MPD contributed to study design, data management, data analysis, data interpretation, and draft of the manuscript. HS and EG contributed to study design, data interpretation and manuscript editing. ABN, KKP, MR, MSA, HMA, MZR, MEH, RCP, SPL, led and participated in site data collection, data interpretation and manuscript reviewing. JV was responsible for laboratory analyses. SC contributed to the data interpretation and manuscript reviewing. All the authors critically reviewed and approved the final version of the manuscript.

## Data sharing

All analyses were performed in R (version 4.2.3). Data and source code to reproduce analyses will be available at https://github.com/marianaperezduque.

## Acknowledgements

This study was funded by the CDC, MPD is funded by the Gates Cambridge Trust (OPP1144), HS was funded by the ERC (804744), SC acknowledges financial support from Labeix IBEID (Grant ANR-10-LABX-62-IBEID), the European Union’s Horizon 2020 research and innovation programme under VEO grant agreement (874735)

---

**Supplementary Figure 1.**
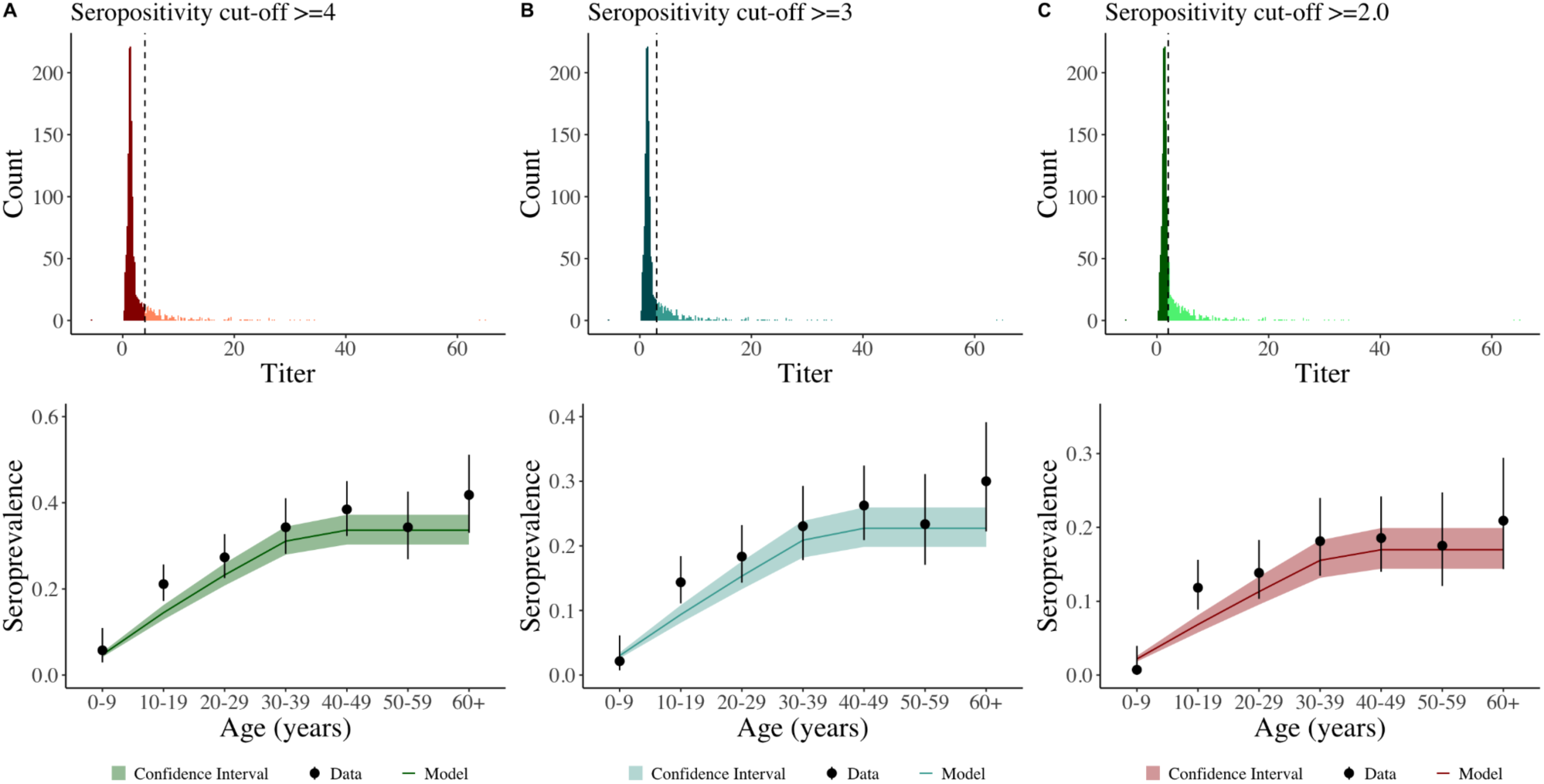
Sensitivity analysis around JEV titer cut point. Titer is the relative median fluorescence intensity value, obtained dividing the median fluorescence intensity measured over a background control.

**Supplementary Figure 2.**
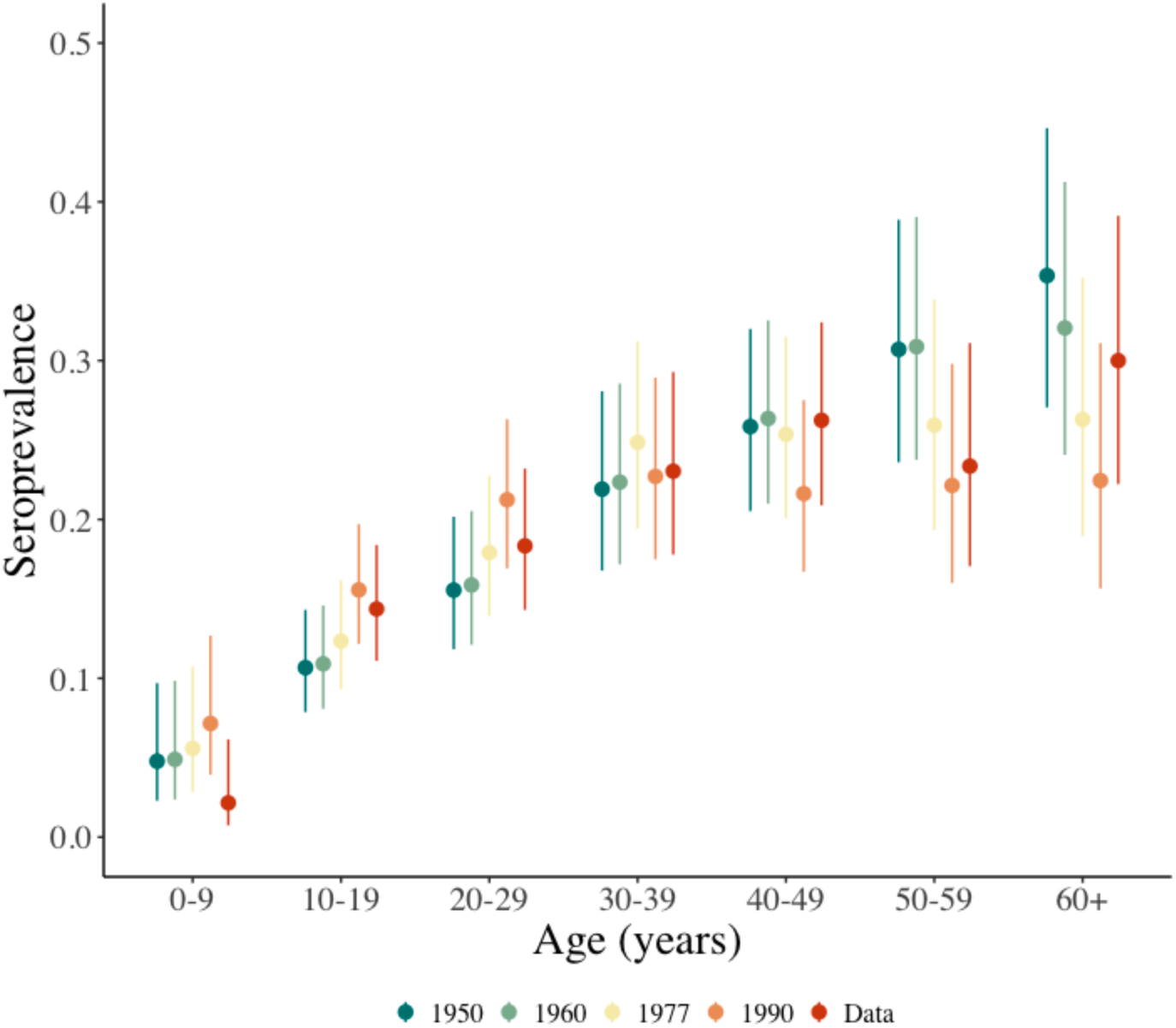
Sensitivity analysis around JEV introduction year. Figure shows JEV IgG seroprevalence and 95%CI (point and range) by age group and coloured by JEV introduction year (1950, 1960, 1977, 1990).

**Supplementary Figure 3.**
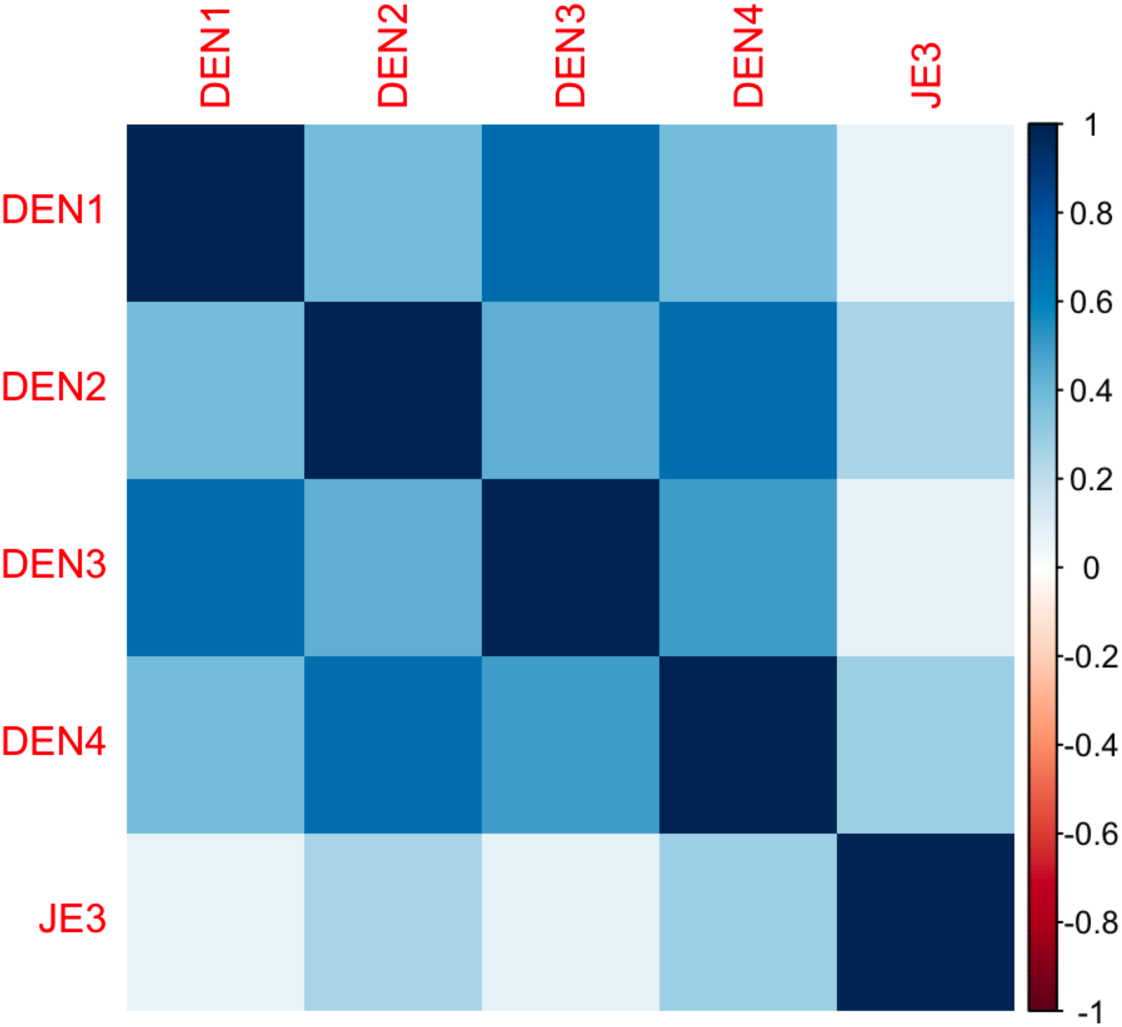
Mean relative fluorescence intensity of the JEV antigen with the mean relative fluorescence intensity to the four DENV1-4 antigens.

**Supplementary Figure 4.**
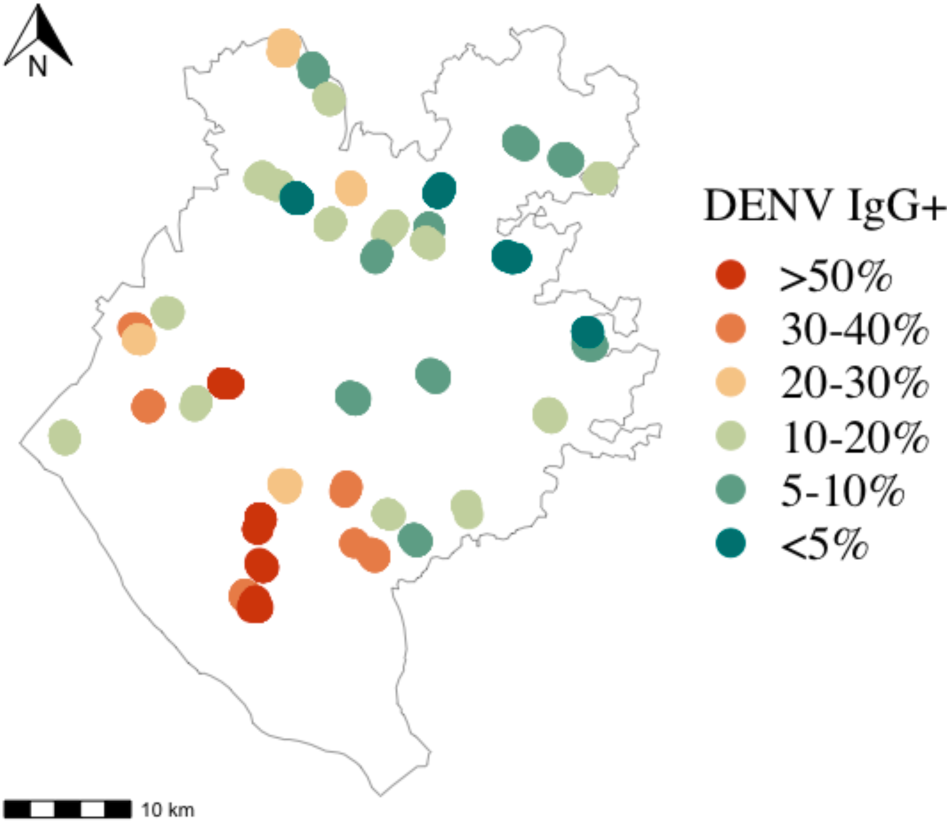
Map of sampled communities locations in Chapai Nawabganj and proportion of human individuals of DENV IgG seropositive.

**Supplementary Table 1.**
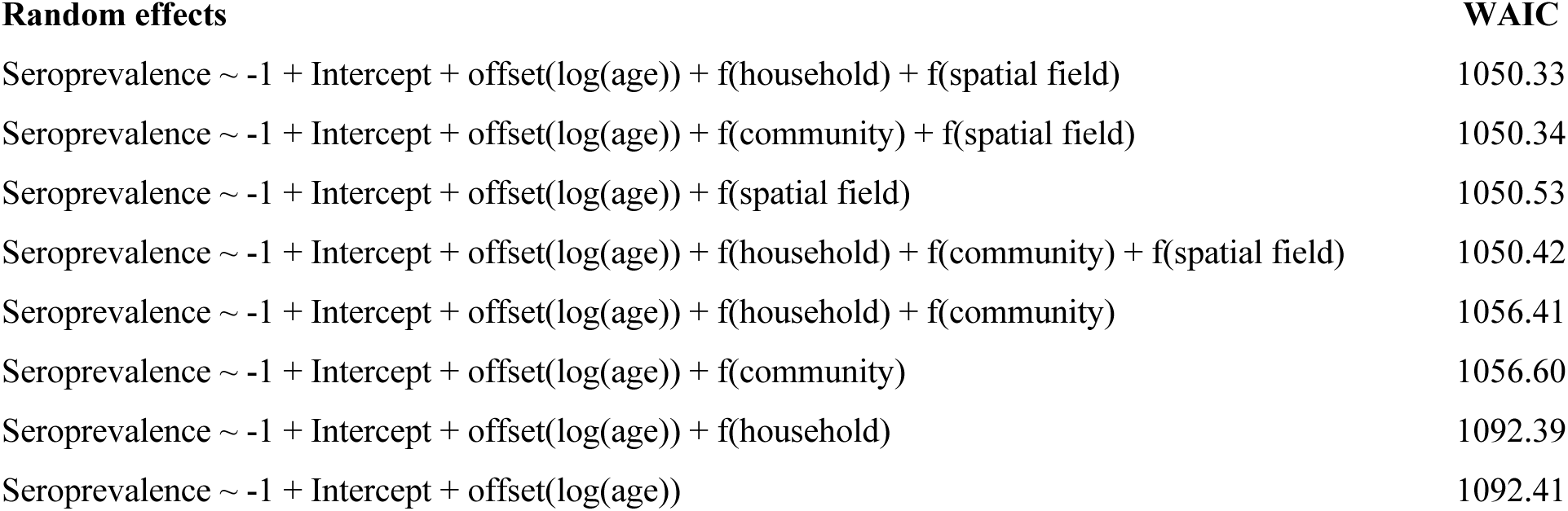
Mixed effects multivariate logistic regression models and corresponding Widely Applicable Bayesian Information Criterion (WAIC).

